# EXPLORING CLINICIANS’ PERSPECTIVES TOWARDS AI-RADIOLOGY & ITS CLINICAL ADOPTION: A QUALITATIVE STUDY FROM PAKISTAN

**DOI:** 10.64898/2026.02.26.26347151

**Authors:** Iqra Bismillah, Shiyam S. Tikmani, Shaista Afzal, Nasreen Naz, Lubna B Vohra

## Abstract

AI is already finding its way into the diagnostic radiology realm of various regions around the world, but there is still a lack of evidence on the situation in LMICs. This qualitative study examined the research problem through the perspectives of clinicians regarding the adoption of AI-Radiology in Karachi, Pakistan, using the Technology Acceptance Model and interpreting the results into practice and policy using the Problem Driven Iteration Adaptation lens. 13 clinicians (radiologists, tertiary care hospitals) were interviewed between May and August 2025. The semi structured interviews were audio recorded, transcribed and coded in NVivo 14. TAM constructs of perceived usefulness and perceived ease of use were analyzed in a deductive content analysis, and interpretation of implementation pathways was informed by PDIA. Four themes emerged. Implementation attitudes were realistic optimism. The subjects put AI in terms of an assistant and second reader, and clinical judgment and accountability could not be delegated. Issues centered on privacy of data, and over dependence. Perceived ease of use was based on training, infrastructure, fit in workflow and trust. Costs, poor connectivity, the lack of institutional capacity, and generational resistance were the barriers whereas triage acceleration, mass screening support, workload reduction, and time saving were the facilitators. For adoption, education, practical upskilling, guidelines, and local clinical approval were requirements. The greatest perceived usefulness was in situations where AI was applied to specific bottlenecks like quick screening, quantitative measurements, remote-area reporting, and trainees’ decision support; the constraints included data quality, generalizability, and algorithm error, the risk of confidentiality, and the impossibility to substitute contextual clinical reasoning. Such priorities as national and institutional data protection policies, formal vetting of tools, smooth integration with radiology information systems and AI literacy in the curriculum were included. The sample is limited to one city and the qualitative design does not enhance generalizability but the results provide practical recommendations. The mixed resource setting of Karachi is a potential place where AI can be a reliable collaborator in the field of radiology in case of adequate infrastructure and training of clinicians and a long-term human control. Perceived usefulness can be converted to routine and safe clinical use with strategic and staged implementation.

## 2. INTRODUCTION

### 2.1 Background & Context

#### 2.1.1 Artificial Intelligence

A computerized altered system that is capable of the work that is similar to the human intelligence and is mainly problem solving, decision making and command understanding (1). A computerized altered system that is capable of the work that is similar to the human intelligence and is mainly problem solving, decision making and command understanding

#### 2.1.2 AI in Healthcare

AI is a new area that has found use in health facilities in different ways (2). The AI models will be able to do a general scope of work such as the one where doctors assist in clinical decision making, treatment protocols and task prioritization. The development of AI has followed progress in computing capabilities and the access to bigger data sets, which speed up the development of AI and opens a possibility to enhance the quality of efficiency and access to health services with the potential of cost reduction. Nevertheless, the effective implementation of the AI in the healthcare systems does not only rely on the technical performance, but also on the acceptance governance frameworks and compatibility with the already existing clinical workflow by the clinicians.

#### 2.1.3 AI in Radiology

The field of medical imaging can be considered among the most dynamic areas of AI implementation because it is based on massively organized visual information and pattern recognition. The application of AI based tools is currently applied to imaging mortalities such as X-ray, CT, MRI, Ultrasound and mammography to detect the disease, to segment the image and to identify anomalies and to stratify the rest. The implications of these developments on the role of radiologists, clinical accountability and diagnostic decision making have been noted. Consequently, AI radiology has become the most important field on health system innovation especially where substituting workforce is increasingly challenged and the increasing diagnostic needs. (3) (4).

### 2.2 Problem Statement

Although global initiatives to implement AI in healthcare have been successful and applied in Pakistan, the rate of adoption is low and uneven, and interest in AI-assisted diagnostic technology is increasing despite the absence of qualitative research on clinician attitude towards AI-radiology and the situation in low-and middle-income countries (LMIC) settings. Available literature concentrates tremendously on technological performance or on high-income conditions with minimal consideration on clinician beliefs issues and expectations. Unless these perspectives are comprehended, AI implementation may have a low uptake, improper utilization and ineffective resource distribution that may compromise health system objectives. (3)

### 2.3 Rationale

Awareness, beliefs, and intentions of clinicians to implement AI could be used to devise focused interventions to address resistance and subsequent adoption. As the largest metropolitan city in Pakistan, Karachi can be used as a representative to understand the views of the clinicians because of the dynamic nature of healthcare in the city (5). The research presents an important awareness gap, which will be used to make informed policies to develop AI implementation strategies specific to the context of Pakistan healthcare.

### 2.4 Objective

In an attempt to understand the clinicians and radiologists awareness, their views and attitudes towards AI assisted diagnostic Radiology and its introduction into clinical practice in Karachi Pakistan.

### 2.5 Research Questions

2.5.1 Which factors will affect the acceptance or rejection of clinicians to utilize AI-Radiology to diagnose the disease?

2.5.2 What does the views of the clinicians and radiologists on AI-Radiology tell the policymakers to formulate efficient policies to incorporate AI in Radiology?

## 3. METHODOLOGY

### 3.1 Methods

#### 3.1.1 Study Design

Qualitative descriptive exploratory study design was applied. The paradigm used was constructivism. Our study design is founded on the underlying philosophical assumption of ontological stance of relativism and the assumption that the reality is subjective and multiple with respect to the perceptions and context of individuals.

### 3.2 Conceptual Frameworks

#### 3.2.1 Theoretical (TAM) & Analytical (PDIA) Frameworks

The research has used a multi framework methodology that guaranteed both theoretical and practical relevance. The TAM (6) was used as the main analytical tool in the process of data analysis and PDIA was used to inform the applied phase of providing the research insight and systems or policy level pathways. The integrated conceptual framework is shown in **Error! Reference source not found**..

**Fig 1:**
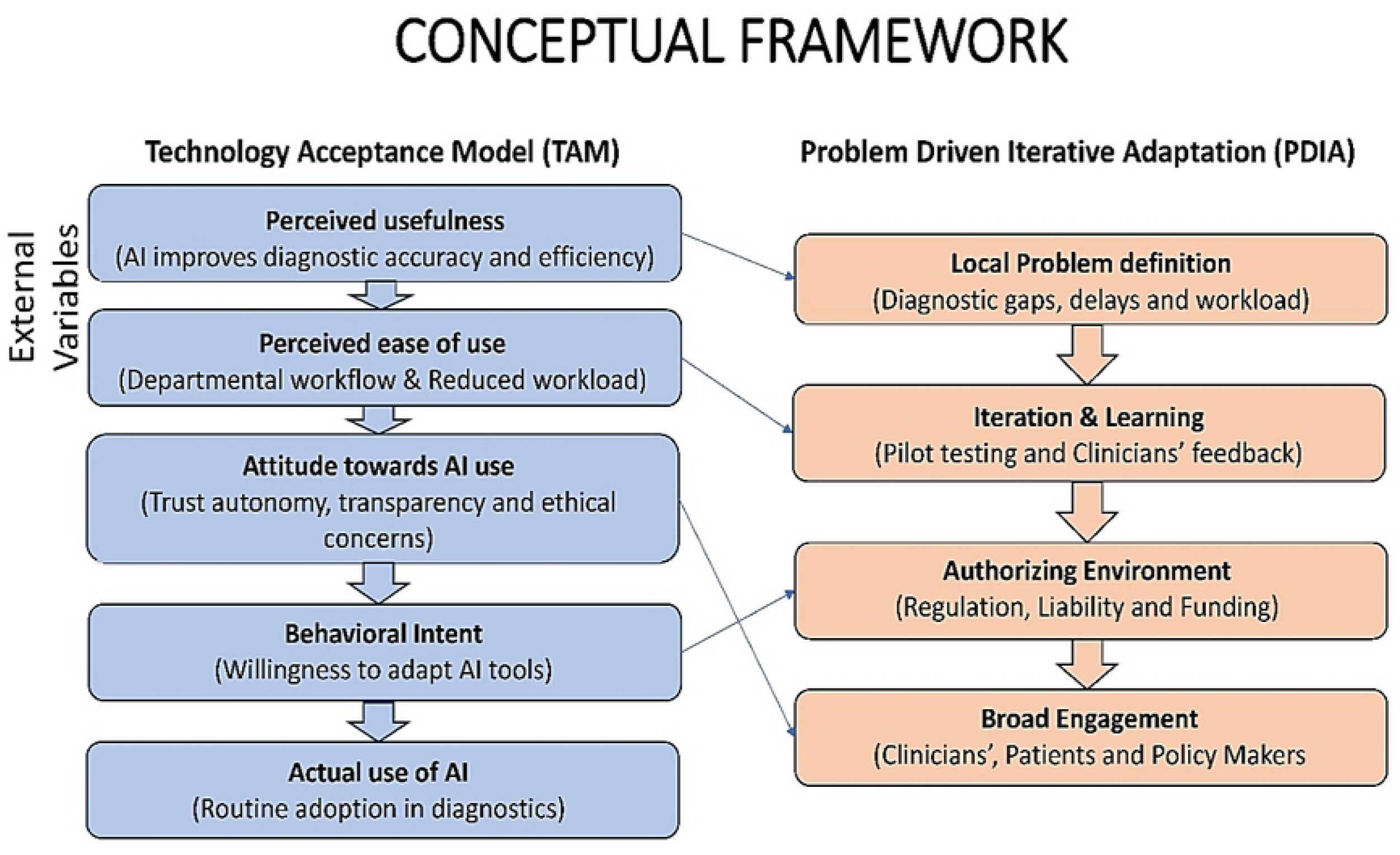
Conceptual framework integrating the TAM and PDIA.

### 3.3 Participants Selection

Online professional forums and networks were used to select 13 clinicians and radiologists. The recruitment period commenced on 15^th^ April, 2025 and concluded on 7^th^ May, 2025. The inclusion criteria were the clinicians to be members of both the public and private tertiary care hospitals, which had functional radiological facilities.

### 3.4 Sample Size

We conducted 13 very intensive interviews with clinicians and radiologists. The concept used to determine this size of sample was that of information power.

### 3.5 Sampling Strategy

Purposive method of sampling was used so that there is diversity in terms of clinical specialties. Table 1 elaborates the recruitment plan, showing the type of participant, how they will be invited, and the reason as to why they will be included (7).

**Table 1:** Recruitment Strategy.

| Discipline | Number of Participants | Recruitment Method | Rationale for Inclusion |
| --- | --- | --- | --- |
| Medicine | 3 | Direct Invitations via email, professional networks, and medical societies | Provide insights on AI integration in personalized medicines and diagnostic practices |
| Surgery | 3 | Direct Invitations, professional networks, and surgical societies | Offer perspectives on AI in surgical imaging and diagnostic tools |
| Gynecology | 1 | Direct Invitations via professional networks, and gynecological societies | Provide viewpoints on AI based diagnosis of obstetric and gynecological imaging |
| Radiology | 3 | Direct invitations to radiologists through radiological societies | Core expertise in AI applications for diagnostic imaging |
| Pediatrics | 3 | Direct invitations via professional networks | Give their insights on AI assisted pediatric radiodiagnosis |

### 3.6 Considerations

Of the 19 physicians contacted in the tertiary care hospitals, 10 clinicians and 3 radiologists accepted to take part in this project. The potential participants were contacted and approached through emails and informed about the purpose and procedure of the study.

### 3.7 Study Settings

The tertiary care hospitals of Karachi were selected to recruit the participants because the city has a full spectrum of public and military hospitals that provide advanced diagnostic and clinical services to the various population with radiological services such as digital X-ray CT, MRI and ultrasound services fully operational.

### 3.8 Data Collection Methods

The semi-structured interview guide was sent to the chosen participants through emails.

#### 3.8.1 Permission & consent

prior to the start of the data collection activity, consent was obtained by each of the participants.

#### 3.8.2 Data collection tools

The data was gathered using English. The interviews were made once and took about 30 minutes. The interviews were carried out using field notes to document the non-verbal data and observations. We used face to face interviews as our medium and the study was carried out between May 21^st^ - August 30^th^, 2025. The interviews were all audio-taped. The audios were transcribed professionally and subsequently, were imported to NVivo to be coded and analyzed.

### 3.9 Data Analysis

Semi structured interview data were analysed by a deductive content analysis that was informed by the major constructs of TAM model. The data was systematically arranged as far as the theoretical constructs were concerned. We have reviewed the transcripts after data cleaning and anonymization, systematic alignment and coding of the data based on research questions based on Technology acceptance model (TAM). The finalized codebook containing themes, subthemes, codes, number of contributing participants, and excerpt counts is provided in Table 2.

**Table 2:** Codebook of Identified themes and narratives.

| Themes | Sub-Themes | Codes | Narratives |
| --- | --- | --- | --- |
| Attitudes Towards Implementation | Critical Implementation Concerns | Clinical Accountability Rests with Doctor | *"AI can help reduce our workload, but it cannot replace the radiologist's eyes and expertise," explained one participant (R03)* |
|  |  | Clinical Judgement is the ultimate check for accountability | *should "always be double-checked by the radiologist" (M01)* |
|  |  | Cybersecurity concern | 1 |
|  |  | Improves Inter-Departmental Communication | 2 |
|  |  | Patient data privacy concern | 1 |
|  |  | Vulnerability to data leakage | 2 |
|  | Overall, Stance and Beliefs | AI as an Assistant, not a Replacement | 6 |
|  |  | AI can be a hindrance | 2 |
|  |  | AI provides everything fake | 1 |
|  |  | Convenient and accuracy | 1 |
|  |  | Double check the references provided by AI | 1 |
|  |  | In Future Everything will be governed by AI | 2 |
|  |  | Not accepting AI as a Doctor | 1 |
|  |  | Patient Trust in Doctor's Tech Concerns | 5 |
|  |  | Positive experience of using it | 4 |
|  |  | Reliance on AI as well as radiologists | 1 |
|  |  | Use of AI is not about patient's choice | 1 |
| Perceived Ease of Use | Barriers to Adoption | Financial Constraints |  |
|  |  | Lack of Awareness and Training | 3 |
|  |  | Lack of Specific Medical Training | *"If we suddenly get an AI software, we'd need to learn from scratch—and there's no structured training available" (P03).* |
|  |  | Lack Training Infrastructure | 4 |
|  |  | Old Age Patients | 4 |
|  |  | Technological and Infrastructural Limitations | *"Stay current and updated with the field" (M02)* |
|  |  | Trust issues with AI | 7 |
|  |  | Weak Connection Network | 1 |
|  | Drivers of Acceptance | Addressing Workforce Shortages | 1 |
|  |  | AI are safer than human beings | 1 |
|  |  | Aids in Mass Screening | *"If AI helps the radiologist not miss a fracture or tumor, the patient gets the correct diagnosis sooner" (P02)* |
|  |  | Better Accessibility | 1 |
|  |  | Diagnostic Aid and Confidence Building | *"AI can pick up subtle findings that a human might miss, especially when we're tired," explained one radiologist (R03)* |
|  |  | Easily accepting to make work easy | 4 |
|  |  | Financially Cost-Effective | 1 |
|  |  | Guidance in Absence of Expertise | 2 |
|  |  | Helps to start treatment on time | 1 |
|  |  | Improved Accessibility in Remote Areas | 4 |
|  |  | Need to specific in prompts | 2 |
|  |  | Time-Saving and Efficient | 4 |
|  |  | Workload Reduction and Burnout Prevention | 8 |
|  | Prerequisites for Adoption | Development of Ethical and Regulatory Guidelines | 5 |
|  |  | Integration into Medical Education | 6 |
|  |  | Knowing How to Use | 6 |
|  |  | Need for Validation and Evidence | 9 |
|  |  | Should include basic medical education system | 5 |
|  | Resistance due to perceived disadvantages | AI has no authentic knowledge | 4 |
|  |  | Fear of Job Replacement or Deskilling |  |
|  |  | Generational and Cultural Resistance |  |
|  |  | Human psychology barrier |  |
|  |  | Lack of Trust and Reliability Concerns |  |
|  |  | Too much dependency on AI |  |
| Perceived Usefulness of AI in Radiology | Perceived Advantages | Accuracy and Error Reduction |  |
|  |  | Departmental workflow efficiency |  |
|  |  | Enhanced Learning and Self-Improvement |  |
|  |  | Learning and Reference Tool |  |
|  |  | Positive impact on radiology |  |
|  |  | Remote reporting improves access |  |
|  |  | Save the patient from being undergone surgical intervention |  |
|  |  | Screening Programs |  |
|  |  | Speed and Efficiency in Diagnosis |  |
|  |  | To stay current and updated |  |
|  |  | Triage and Prioritization |  |
|  |  | Triage Efficiency |  |
|  |  | Use of AI like ChatGPT, NoteKL for literature analyses |  |
|  |  | Used in routine practices |  |
|  | Perceived Limitations and Risks | Confidentiality risk in cloud data |  |
|  |  | Data privacy concern |  |
|  |  | Dependence on Data Quality and Algorithms |  |
|  |  | Hallucinations and Fabricated Information |  |
|  |  | Inability to Replace Clinical Judgment |  |
|  |  | No privacy on internet |  |
|  |  | Not Self-Sufficient for Diagnosis |  |
|  |  | People from old era concerns |  |
|  |  | Potential for Incorrect or Outdated Information |  |
|  |  | Unusual Cases |  |
| Recommendation and policy practice | Policies or practical change | Avoid saving data |  |
|  |  | Ethical Concerns Issues | *"We need our health authorities to set protocols—who is responsible for errors and* *how to validate an AI model,” urged a senior radiologist (R03)* |
|  |  | Inform patient about using the technique | 2 |
|  |  | Need of vetting program | *“Before an AI is used clinically, there should be a vetting program or pilot approval,” explained a clinician (S03).* |
|  |  | No Major Ethical Concerns in Radiology |  |
|  |  | Policies and Public awareness |  |
|  |  | Presence of Ethical committee |  |
|  | Recommendations | Accuracy improvement |  |
|  |  | Adoption is necessary in this changing world |  |
|  |  | AI in Syllabus |  |
|  |  | Important to Understand Radiology |  |
|  |  | Integrate with AI-assisted radio diagnosis in task shifting areas | *“Hospitals should have rules against saving sensitive data unless encrypted and absolutely necessary,” noted one clinician (S03).* |
|  |  | Save Patients Surgical Intervention | 5 |
|  |  | Training Programs Doctors | *“There should be workshops for clinicians on how to use and interpret AI tools,” said one clinician (G01).* |

### 3.10 Rigor

One method of eliminating reporting bias was based on (COREQ) 32-item checklist (8). These data were stored in a safe file that was password-protected and only accessible by the authorized officials.

### 3.11 Credibility

Triangulation of data source was obtained through gathering the knowledge of clinicians with various specialties. Triangulation was done theoretically through the application of TAM as an analytical framework. Triage Triangulation Investigator Triangulation It was performed by the effort of the research team reviewing NVivo 14 refining the interpretations and improving analytic rigor.

To achieve that the categories and codes were illustrated by the use of quotations of authentic participants (9).

### 3.12 Trustworthiness

The interviews were studied by our team of researchers several times in order to enhance the credibility.

### 3.13 Dependability

After the process of data collection and data analysis that was systematically based on the TAMs and under the influence of NVivo 14 software that ensured the explicit coding and recording of our analytical choices.

### 3.14 Confirmability

An audit trail was also kept which involved recording of interview transcripts, field notes, coding structures and analysis memos as a transparency.

### 3.15 Transferability

The exact description of the level of experience of the participants enabled the reader to gauge the degree of similarity of the context, as well as the possibility of other researchers or policy makers to ascertain the generalizability to other health care situations in LMICs.

### 3.16 Reflexive Statement / Reflexivity

The assumptions were recognised to counterbalance its effects on data interpretation. In order to prevent such biases reflexive journals were kept and Peer reviews were done.

### 3.17 Ethical considerations

ERC of Aga Khan University (Ref No. 2025-11180-34032) and National Bioethics Committee approved Research, National Institute of Health, Pakistan (Ref No.4-87/NBCR-1290/24-25/380).

## 4. RESULTS

### 4.1 Findings

This had 13 clinicians involved. The majority were familiar with the fundamental idea of AI-Radiology, but some of them were direct users of AI tools. The summary of the demographics of the participants is presented in Table 3.

**Table 3:**
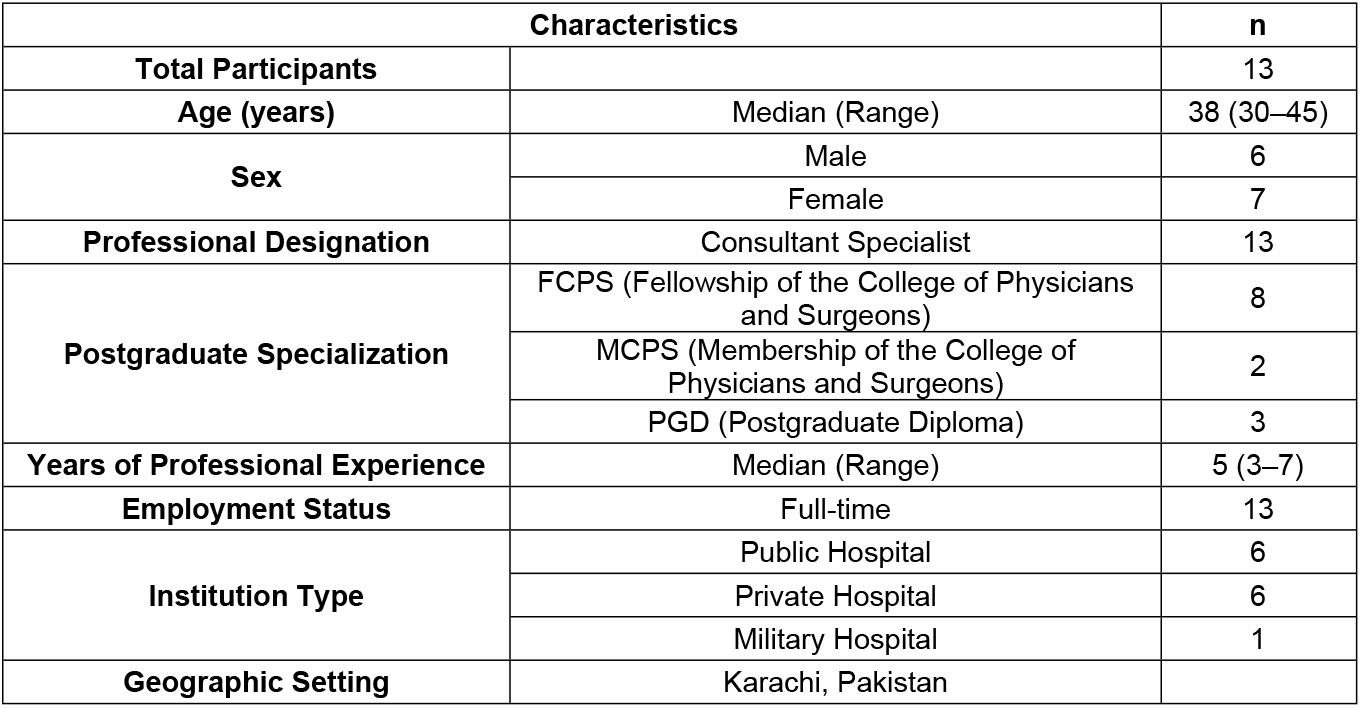
Summary of clinicians’ demographic and professional characteristics included in the study.

| Characteristics |  | n |
| --- | --- | --- |
| Total Participants |  | 13 |
| Age (years) | Median (Range) | 38 (30–45) |
| Sex | Male | 6 |
|  | Female | 7 |
| Professional Designation | Consultant Specialist | 13 |
| Postgraduate Specialization | FCPS (Fellowship of the College of Physicians and Surgeons) | 8 |
|  | MCPS (Membership of the College of Physicians and Surgeons) | 2 |
|  | PGD (Postgraduate Diploma) | 3 |
| Years of Professional Experience | Median (Range) | 5 (3–7) |
| Employment Status | Full-time | 13 |
| Institution Type | Public Hospital | 6 |
|  | Private Hospital | 6 |
|  | Military Hospital | 1 |
| Geographic Setting | Karachi, Pakistan |  |

### 4.2 Participants Characteristics

Each theme is presented below with its subthemes and supported by exemplar quotations from participants (quoted participants are identified by their field and number of interviews, e.g., “S01 for Interviewer 1 who is surgeon”). Although this is a qualitative study, Table 3 summarizes participant characteristics and Table 2 reports code frequencies and contributor counts to indicate which issues were most widely raised across interviews. These counts are descriptive and are not interpreted as prevalence estimates.

Using contributor counts as a descriptive signal (Table 2), the most widely raised cross-cutting issues were the non-replaceability of clinical judgement (10 participants), the need for validation/evidence (9 participants), and trust/reliability concerns (9 participants). These directly map to the study objectives by distinguishing (i) core adoption barriers (trust, validation, accountability) from (ii) perceived benefits that motivate intention to use (efficiency and workload reduction).

## 5. DISCUSSION

According to the overall thematic analysis of the perceptions of healthcare professionals towards using Artificial Intelligence (AI) in radiology, the results indicate that there is a complicated interplay of reserved optimism, major practical issues, and a definite picture of what should be done in order to move toward integration. The discussion, which is organized into four overarching themes based on the “Technology Acceptance Model (TAM)” and “Problem-Driven Iterative Adaptation (PDIA) framework”, offers a more detailed insight into the factors affecting AI adoption in this particular clinical scenario. The findings are categorized into four overarching themes, these include “Attitude towards Implementation, Perceived Ease of Use, Perceived Usefulness of AI in Radiology, and Recommendation and Policy Practice”.

### 5.1 The first theme

“Attitudes Towards Implementation”, summarizes a range of sentiments of clinicians, which went both towards optimistic acceptance to entrenched fear. The hierarchy chart and mind map of this theme demonstrate that the essence of the position of clinicians is that AI should act as the helper and not as a substitution, which is deeply rooted in most of the participants. Graphically, it is illustrated by the fact that the theme of AI as an Assistant and Not a Replacement occupies the middle of the thematic structure. The issues around this center are the pivotal concerns about accountability and clinical judgment, which are presented as uncompromising precautions. The comparative chart with its visuals also shows that the positive experiences and the possibility of the enhanced inter-departmental communication also exist, but they are heavily overshadowed by the sense of constant apprehension of data privacy, cybersecurity, and patient trust. The characters are all used to illustrate an approach that is open to technological development but which is essentially conditional on retaining the supreme human control and moral accountability.

### 5.2 The second theme

“Perceived Ease of Use” is graphically subdivided into drivers of and barriers to adoption. The hierarchy chart shows a sharp line of difference existing between the projected benefits and the reality on the ground. On the one hand, workload-reduction, time-saving efficiency, and support in mass screening are among the leading drivers, which implies that there is a high awareness of the possible role of AI in relieving systemic pressures. The charts, on the other hand, demonstrate that the route to these benefits is obstructed by one significant hurdle: a lack of trust and reliability concerns is the most repeated sub-theme. The other significant infrastructural and technological limitations that accompany this are lack of good network connections and lack of a training infrastructure. This mind map was an excellent display of the relationship between these barriers to each other to create a web of achievable barriers that currently make AI a hard thing to implement in the real world without some issues. The comparative chart shows that the technology constraints and trust issues in the data is way higher than the drivers and that is the reason why the adoption rate is low.

### 5.3 Its third theme

“Perceived Usefulness of AI in Radiology”, is graphically arranged in a way that shows the opposition between the enormous benefits and the vital constraints of AI. The hierarchy chart gives it a massive focus to the benefits, and such sub-themes as “Speed and Efficiency in Diagnosis,” “Enhanced Learning and Self-Improvement,” and “Use in Routine Practices” are among the top results with significant participation. This means that there is general acceptance of the usefulness of AI as a diagnostic tool, triage tool, and learning tool. This however is offset by the mind map and the comparative chart which place equal or even greater visual weight on the limitations. The most severe shortcoming, which is mentioned by all participants, is the “Inability to Replace Clinical Judgment”. Another key issue like the possibility of “Incorrect or Outdated Information” and the possibility of hallucinations are also highly visible.

### 5.4 Last is the fourth theme

“Recommendations and Policy Practice” which represents the recommended course of action. The calls of structural and educational reforms prevailed in the hierarchy chart of this theme. The two recommendations that have the most powerful recommendations, based on the evidence, are the “Need for Validation and Evidence” and “the Integration into Medical Education”. The figure further demonstrate that the participants do not only criticize the current state of things but also offer solutions: building of the ethical guidelines, formal training programs and policy of creating the awareness of the citizens are all the brightest points of the mind map. This theme is the ultimate synthesis of the first three themes discussed before, namely, optimistic attitudes, perceived usefulness, and ease of use can be fully achieved only by conscious policy action and structural adjustment.

### 5.5 Recommendation for Adoption and Capacity Building

Most of the participants in the study agreed with the strategic implementation of AI in diagnostic radiology because they believe it is a requirement. In addition, some respondents have stated that it is a necessary step in this changing world because the AI-based technological solution will assist in improving the efficiency and accuracy of clinical processes. Moreover, in case of a dense flow of patients, or the lack of specialists, AI may assist in achieving a more accurate diagnosis.

The participants believed that it would be a good idea to initiate training programs to train doctors with the use of AI because they believe it will demonstrate confidence, ensure the safe introduction of AI, and put the practice of the doctor in alignment with the existing international standards.

The management of AI in radiology through regulation, education, ethical control, and community involvement should be addressed as a catching net to preserve the reliability in the process as regulation, education, ethical control, and social interaction keep the policy’s purpose intact to achieve success and enhance the health system of Pakistan.

### 5.6 Strengths

The research paper has helped in a fairly uncharted field in Pakistan and the LMIC setting at large regarding clinicians attitude towards AI-Radiology. The study presents locally based evidence by covering the policy and ethical aspects of the AI implementation to guide national digital health strategies training programs and ethical governance systems. The practical implications and the recommendations on the study that are driven by the PDI framework direct the practical aspects of the study that can be used to drive the research findings forward not only by understanding the healthcare administration system, but also by providing practical insights to the healthcare administrator, regulation and policy makers to enhance responsible and appropriate AI integration.

### 5.7 Limitations

The sample was rather small that, despite the fact that it is suitable in case of qualitative inquiry and supported by the principles of information power and data saturation, reduces the applicability of the results to the large population of clinicians. The obtained insights are specific to the context of the experiences and the worldviews of clinicians who are working in Karachi and might not accurately depict the differences in other parts of Pakistan or even the differences of other levels of health care settings like rural or secondary facilities. Secondly the deductive analysis based on the TAM model was useful to inform the deductive analysis that may not fully explain all the contextual and systemic factors of AI adoption in LMIC environments.

### 5.8 Study implications

Policymaking wise, this study highlights the importance of national policies and regulations. Respondents made numerous calls to regulate data, establish strict rules of responsibility, and ethical standards prior to the mass use of AI tools. Our results imply that there is a need of certain healthcare provisions. To illustrate the example, regulatory authorities might introduce certification requirements to clinical AI, which will help to certify any tool under the validation of local requirements. Simultaneously, its application should be made possible by government and hospital leadership investing in the proper infrastructure, such as the funding of enterprise-level PACS/IT systems. Lastly, the stakeholders must promote the idea of a public-private collaboration: the experience of the private sector may be involved in developing economical AI-based methods of the approaches to the health system of Pakistan, which are promoted by international radiology projects (10, 11).

### 5.9 Summary

The thematic analysis showed that there was a complex relationship of optimism and caution among the clinicians on AI in the field of radiology.

To be effective in adopting it, education, validation and governance were brought out as key requirements. The respondents proposed moral codes, on-site testing, inclusion of AI in the medical curriculum and national data protection.

In general, clinicians in Karachi agree and feel that AI in Radiology can be a game changer. Nonetheless, they believe that its usage is conditional under the condition of its validation, IT infrastructure and adequacy of ethical transparency. The future of artificial intelligence will not be the replacement of the clinicians or radiologists, but just enhancing them by building trust, competency, and responsibility.

## 6. CONCLUSION

The resulting thematic analysis that is interpreted as the visual representations of the thematic analysis forms the image of the medical community that is at the crossroads. Clinicians in Karachi know the potential of AI in the radiology department in the field of medicine to change their life, efficiency, burn out and precision of the diagnosis. However, the three-fold set of significant challenges such as ingrained problems of mistrust and trust coupled with critical structural and technological bottlenecks and an absence of proper regulatory and educative framework are holding this promise at bay. The outcome will clearly indicate that no successful integration will be done with the implementation of technology itself. It will entail a multi-dimensional strategy that will entail certifying AI as trustworthy through rigorous local testing, investing on the digital infrastructure, introducing massive training programs to existing and upcoming clinicians, and establishing straightforward ethics-related guidelines to control the use and ensure patient data privacy. And, after all, AI has not been seen as a substitute to the radiologist, but as a partner, the work with which he/she should be obtained in a well-considered way, in mutual trust, evidence, and joint responsibility.

## Data Availability

All relevant data are within the manuscript and its Supporting Information files.

